# Serotype patterns of pneumococcal disease in adults are correlated with carriage patterns in older children

**DOI:** 10.1101/2019.12.18.19015180

**Authors:** Anne L. Wyllie, Joshua L. Warren, Gili Regev-Yochay, Noga Givon-Lavi, Ron Dagan, Daniel M. Weinberger

## Abstract

**Background:** The importance of specific serotypes causing invasive pneumococcal disease (IPD) differs by age. Data on pneumococcal carriage in different age groups, along with data on serotype-specific invasiveness, could help to explain these age-related patterns and their implications for vaccination.

**Methods:** Using pneumococcal carriage and disease data from Israel, we evaluated the association between serotype-specific IPD in adults and serotype-specific carriage prevalence among children in different age categories, while adjusting for serotype-specific invasiveness. We used a sliding window approach to estimate carriage prevalence using different age groupings. Deviance Information Criterion was used to determine which age groupings of carriage data best fit the adult IPD data. Serotype-specific disease patterns were further evaluated by stratifying IPD data by comorbidity status.

**Results:** The relative frequency of serotypes causing IPD differed between adults and children, and also differed between older and younger adults and between adults with and without comorbidities. Serotypes over-represented as causes of IPD in adults were more commonly carried in older children as compared to younger children. In line with this, the serotype-specific frequency of carriage in older children (aged 36-59 months), rather than infants, best correlated with serotype-specific IPD in adults.

**Conclusions:** These analyses suggest that older children, rather than infants, are the main drivers of disease patterns in adults. These insights could help in optimizing vaccination strategies to reduce disease burden across all ages.

**40-word summary of the article’s main point:** Serotype-specific rates of invasive pneumococcal disease in adults are better correlated with serotype-specific carriage patterns in older children (36-59 months of age) than those in infants.

## INTRODUCTION

*Streptococcus pneumoniae* (pneumococcus) is a frequent colonizer of the upper respiratory tract of healthy children and is also a major cause of disease globally. The burden of pneumococcal disease disproportionally affects infants and the elderly and those with certain underlying comorbidities [1]. Pneumococcal conjugate vaccines (PCVs) provide protection against 10 or 13 of the 90+ immunologically-distinct polysaccharide capsular types (serotypes), preventing disease and reducing colonization of the nasopharynx in vaccinated individuals. Since children who carry pneumococcus are the main source of exposure for adults [2–4], vaccinating children with PCVs has also resulted in the near-elimination of invasive pneumococcal disease (IPD) caused by vaccine-targeted serotypes (VTs) among adults [5].

Even before the introduction of PCVs, there were important differences in the distribution of serotypes causing IPD between children and adults [6]. For instance, VTs caused a larger fraction of IPD in children. After introduction of PCVs, non-vaccine serotypes (NVTs) have increased in frequency among colonized children. This “serotype replacement” [7] has had a modest effect on IPD rates in children but has substantially reduced the indirect benefits for adults that result from vaccinating infants. Older adults and those with certain underlying diseases have proven particularly susceptible to disease caused by emerging NVTs [8–10]. In some countries, the increase in the incidence of NVTs in adults has offset declines in the incidence of VTs [11].

Variations in vaccine approach and schedule could have important implications for preventing IPD in adults. Currently, the number and timing of doses used in children differs between countries, as does the use of a booster dose. For instance, the United Kingdom recently decided to move to a reduced-dose schedule of PCVs in infants, where children receive a single priming dose and a single booster dose [12]. For such a strategy to be effective in maintaining indirect protection of unvaccinated individuals, it is important that vaccine-derived protection is maintained among the children responsible for transmission in the population. Thus, identifying those age groups driving transmission is critical. Moreover, these issues are increasingly important as new 15- and 20-valent conjugate vaccines move towards licensure.

It is generally assumed that most exposure to pneumococcus in adults results from contact with children [13]. However, children are not all equally likely to carry and transmit pneumococcus, and some groups of children (e.g., preschoolers) might be more influential due to different contact patterns and intensity of carriage [13–15]. Recent work suggests that older children, rather than infants, transmit pneumococcus to adults [13–15].

In this study, we sought to understand drivers of the variations in the distribution of serotypes causing IPD by age and in those with and without comorbidities. We evaluated variations in the serotype distribution in IPD by age and comorbidity, and we tested which age groups of children had serotype patterns in carriage that were most closely correlated with IPD rates in adults. These analyses could help in optimizing vaccine strategies to reduce pneumococcal disease across all age groups.

## METHODS

### Data sources

PCV7 was introduced in Israel in July 2009 using a 2+1 schedule with a catch-up campaign for children <24 months of age. PCV7 was replaced in the schedule by PCV13 starting in November 2010 (without a catch-up). Details of the carriage and IPD data used in this study have been previously described [14,16,17]. Briefly, the carriage data were collected from children visiting the emergency department (ED) at Soroka University Medical Center, the only emergency department in southern Israel. The first 4 Jewish and first 4 Bedouin children <5 years of age visiting the ED each day for any complaint were enrolled in the study to obtain a nasopharyngeal swab. IPD data were obtained from a nationwide surveillance system [16,18]. The carriage and IPD studies were approved by the Sheba Medical Center and the Soroka University Medical Center Institutional Review Boards (IRB). Data in the current study was de-identified before analysis.

Carriage and disease data were divided into 4 equal time periods (Early-PCV7: November 2009 to July 2011, Early PCV13: August 2011 to February 2013, Late-PCV13: March 2013 to October 2014, Stable-Post-PCV period: November 2014 to June 2016). IPD data from 4,304 individuals were stratified by age group (<5 [n=1,180], 5-17 [n=299], 18-39 [n=407], 40-64, [n=943] 65-79 [n=816], 80+ [n=658] years) and according to comorbidity status (no risk, at risk, or high risk for pneumococcal disease [18]) based on recommendations for receipt of PPV23. Risk group analyses were conducted on individuals aged 18+ years who had information on comorbidity status, serotype, and sample collection date. Period 4 for the subset of data with comorbidities ended December 2015 (as compared to June 2016 for the full dataset).

The carriage data were stratified by both ethnicity (Jewish versus Bedouin) and according to presence or absence of recorded complaints that could be caused by pneumococcus (bacteremia, conjunctivitis, influenza, LRI, meningitis, otitis media, pneumonia, sepsis, URI) to confirm results were not sensitive to these factors.

### Modeling the relationship between carriage in children and invasive disease in adults

The goal for these analyses was (1) to evaluate variations in serotype-specific IPD incidence by age and (2) to determine whether carriage data from specific age groups better correlated with IPD incidence. To do this, we used a previously-described Poisson regression model [19]. The outcome variable was the number of IPD cases caused by each serotype in the relevant age group and time period. The covariates in this initial model were the serotype-specific carriage prevalence in the corresponding time period in Jewish children <5 years of age (log transformed) and the serotype-specific invasiveness for children (log-transformed). A serotype-specific random intercept was included to account for unexplained serotype-specific variations in IPD rates. This random intercept can be interpreted as a log(rate ratio) which indicates how much more or less disease was caused by that serotype in adults based on the carriage prevalence in children and invasiveness patterns in children. Analyses were stratified by age or by co-morbidity status. To account for uncertainty in the predictors (serotype-specific carriage and invasiveness when sparse serotype-specific data is available), we fit this model within a Bayesian framework. Further details on the model structure and the priors are reported in [19].

Serotype-specific invasiveness estimates for children <5 years of age, and their uncertainty, were estimated using a separate Bayesian statistical model [19]. Invasiveness was calculated by dividing serotype-specific IPD incidence by the serotype-specific carriage prevalence in <5 year olds [20]. The hierarchical structure of this model effectively allowed us to obtain an estimate of invasiveness (with uncertainty) even for serotypes with few cases of disease or few carriers. The models were fit using JAGS v4.2.0 [21] in RStudio v1.0.143 [22], using R v3.5.1 (https://www.r-project.org/).

Because the IPD data were primarily available for the Jewish population, only carriage and IPD data from Jewish residents were used in the subsequent analyses.

### Stratification of carriage in children by age group to better explain adult IPD patterns

The goal for this analysis was to determine whether serotype-specific carriage prevalence in any particular age group better explained patterns of IPD observed in adults and in different risk groups. To accomplish this, the carriage data from Jewish children were further stratified by age category (≤12 months, <18 months, <24 months, <36 months, 13-59 months, 18-59 months, 24-59 months, and 36-59 months; **Table 1**). The models of IPD described above were re-fit using these subsets of carriage prevalence data. Deviance Information Criteria (DIC) was computed for each model [23]. This Bayesian model comparison metric accounts for the fit of the model to the data while penalizing more complex models. A lower DIC value for a model among a group of competitors indicates that it has an improved balance between model fit and complexity.

**Table 1.** Characteristics of the Data

| Table 1. Characteristics of the Data |  |  |
| --- | --- | --- |
|  | Total number of swabs<br>collected, n | Number of swabs positive for<br>pneumococcus, n (%) |
| **All** | **10379** | **4957 (48)** |
| **Jewish** | **4464** | **1900 (43)** |
| <12 months | 1862 | 717 (39) |
| <18 months | 2491 | 1006 (44) |
| <24 months | 3082 | 1295 (43) |
| <36 months | 3757 | 1611 (43) |
| 13-59 months | 2602 | 1183 (45) |
| 18-59 months | 1973 | 894 (45) |
| 24-59 months | 1382 | 605 (44) |
| 36-59 months | 707 | 289 (41) |
| **Bedouin** | **5915** | **3057 (52)** |
| **Respiratory complaint** | **4279** | **2194 (51)** |
| **Without respiratory**<br>**complaint** | **6118** | **2763 (45)** |

## RESULTS

### Characteristics of the data

Nasopharyngeal swabs from 10,379 individuals <5 years of age were collected between November 2009 and June 2016. Of these, 48% tested positive for pneumococcal carriage. Prevalence was higher in Bedouin children and among those presenting with a complaint that could potentially have been caused by pneumococcus (e.g., otitis media, pneumonia) (**Table 1**). Carriage prevalence increased through the first year of life, then stabilized, declining slightly after 48 months of age in both study populations, though more sharply in Jewish children (**Supplementary Figure S1)**. More swabs were collected from younger than older children (**Supplementary Figure S2**). The serotype distribution was broadly similar in pneumococcal-positive swabs obtained from children with respiratory complaints and those obtained from children without respiratory complaints (**Supplementary Figure S3**). There were some differences in serotype distribution between Jewish and Bedouin children (**Supplementary Figure S4**). For instance, serotype 23A was more common in Jewish children in periods 2-4 and 17F more common in Bedouin children.

There were 4,303 cases of IPD among individuals >5 years of age, with 1,474 (34%) ≥65 years of age. Of those aged 18 years and older, 2,347 individuals had a recorded comorbidity status (no risk, at risk, or high risk for pneumococcal disease [18]) based on recommendations for receipt of PPV23

### Variations in serotype-specific disease patterns by age and comorbidity group

For most serotypes, the observed IPD incidence in adults was similar to what would be expected based on the carriage prevalence and invasiveness estimates from children <5 years of age (random intercept close to zero) (**Figure 1**, results for adults aged 80 years and older; **Supplementary Figure S5**, results for all age groups). Some serotypes however, caused more disease in certain age groups compared to what would be expected based on carriage patterns in children (random intercepts above zero). This was notable for serotype 1, which was most over-represented among 5-17 year olds (with similar patterns observed for serotypes 2 and 17F), and serotype 8, which was most represented among 18-39 year olds (**Figure 2**). Both serotypes demonstrated declining gradients with older age, where the number of IPD cases in older age groups was closer to the expected level. In contrast, serotypes 3, 6C, 15A and 31 were over-represented as causes of IPD more among older adults than among younger adults. Conversely, serotypes 21 and 27 were under-represented as causes of disease in all adults >40 years of age.

**Figure 1.**
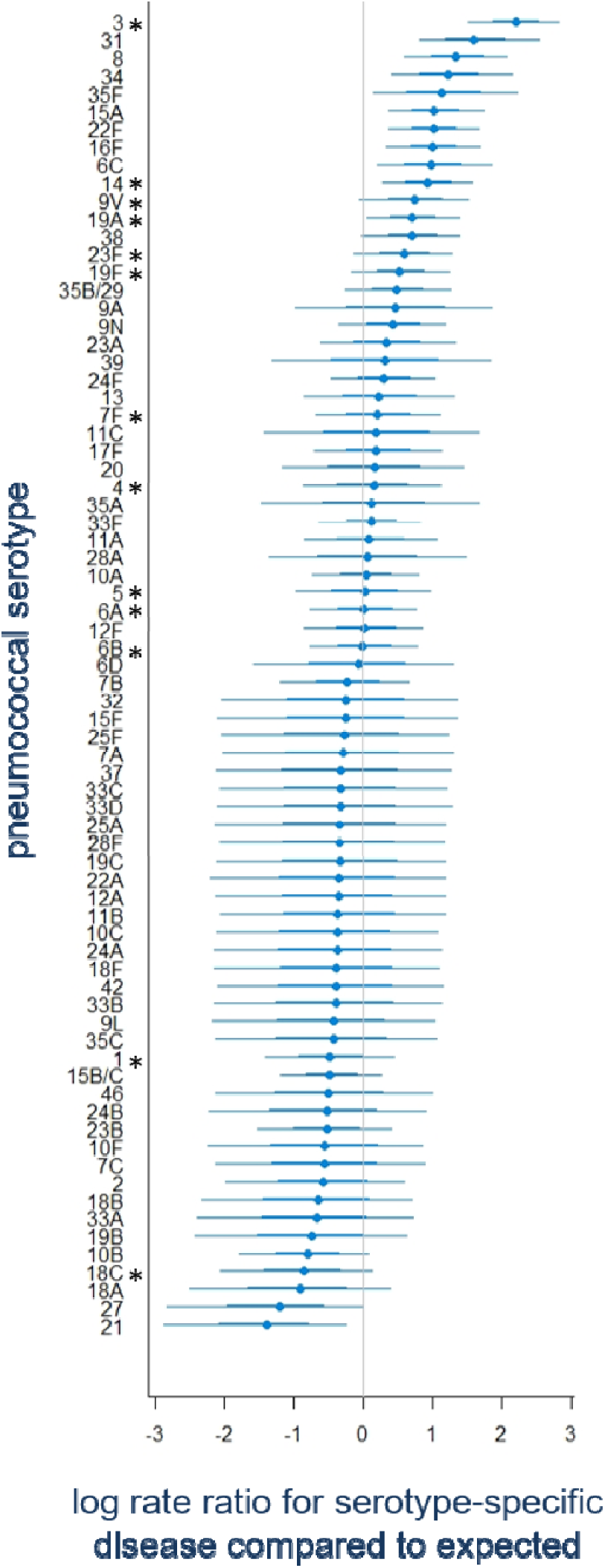
Over-representation of serotypes causing IPD in older adults. The numbers denote serotype-specific random intercepts from a model fit to IPD data from 80+ year old adults in Israel. Values above zero indicate that the serotype is over-represented as a cause of IPD in this age group based on how frequently they are carried in children <5 years of age and their invasiveness in <5 year olds. Values below zero indicate the serotype is under-represented in IPD. For each serotype the 95% (thinner line) and 68% (thicker line) credible intervals are shown. PCV13 vaccine serotypes are denoted by *.

**Figure 2.**
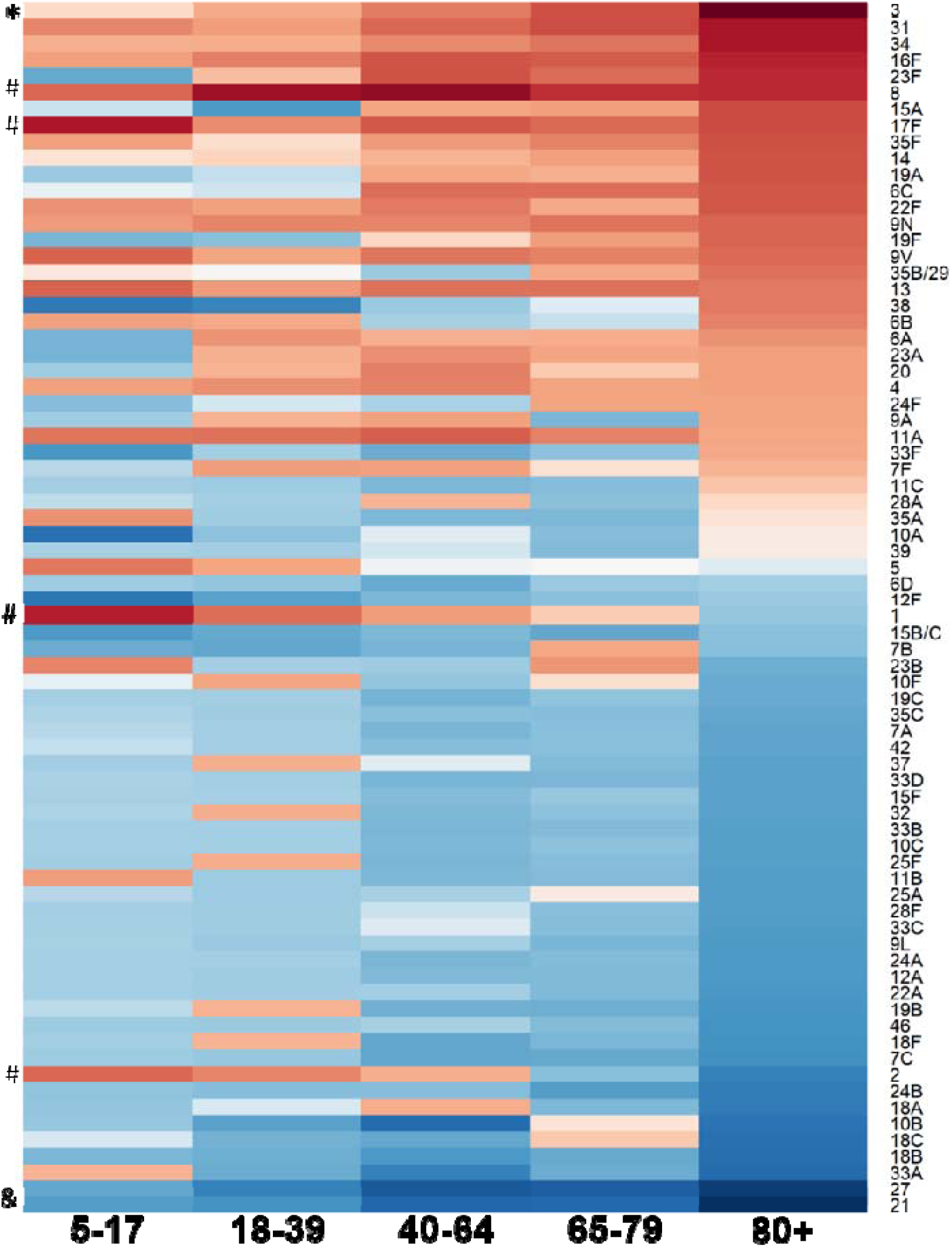
Over-representation of serotypes causing IPD in different age groups. The colors reflect serotype-specific random intercepts from a model fit to IPD data from different age groups in Israel. Darker red represents serotypes in that age strata over-represented as causes of IPD based on how frequently they are carried in children <5 years of age and their invasiveness in <5 year olds. Darker blue indicates serotypes under-represented as causes of IPD in that age group. Symbols on the left highlight age-related patterns for certain serotypes. Those denoted by * become increasingly over-represented with increasing host age, those denoted by & became increasingly under-represented with increasing host age and serotypes denoted by # are more over-represented in younger individuals.

Similar patterns were seen when stratifying by comorbidity status (**Supplementary Figures S6 and S7**). For instance, serotype 3 and 8 were over-represented in adults with and without comorbidities, but the effect was more dramatic in those with comorbid conditions. Likewise, serotype 8 was most notable among those without comorbid conditions. We also considered whether the age or comorbidity patterns were confounding each other. Even after stratifying by comorbidity and by age, some serotypes, and particularly serotype 3, were still over-represented among older adults (**Supplementary Figure S8**).

### Serotype prevalence in carriage differs between younger and older children

We next compared the prevalence of serotypes colonizing children <24 months of age versus those carried by children 24-59 months of age (**Figure 3**). Serotypes 5, 1, 7F, 3, 27, 33A, 6D, 35A, 23F, 14, 34, 18C, 22F, 12A, 24A, 37, 4, 20, 31, 19A were all at least 50% more prevalent in children 24-59 months of age than in those <24months of age. In contrast, serotypes 6A, 10A, 13, 19F, and 21 were at least 25% less prevalent in the children aged 24-35 months.

**Figure 3.**
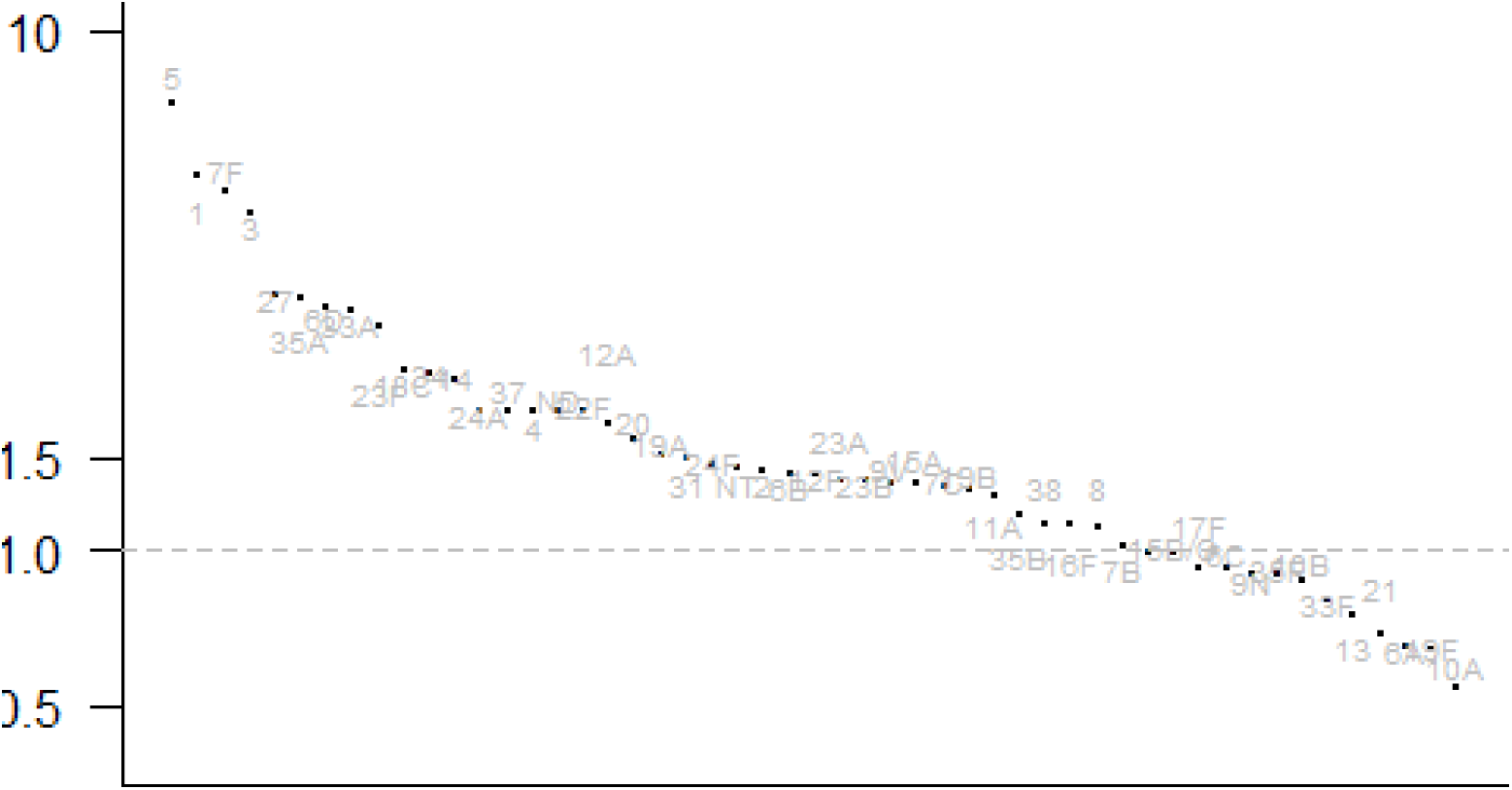
Ratio of serotype-specific carriage prevalence in children aged 24-59 months of age compared to children aged under 24 months.

Comparing the serotypes that were more prevalent in carriage in older versus younger children with the serotypes that were over-represented as causes of IPD in adults, several serotypes at the extremes stand out (**Figure 4)**. In particular, serotype 1 was carried more in the older children than in the younger children and was also among the most over-represented serotypes causing IPD among the 5-17 and 18-39 year old adults. Likewise, serotype 3 was more common in carriage among older children and was over-represented as a cause of disease among adults 40+ years of age.

**Figure 4.**
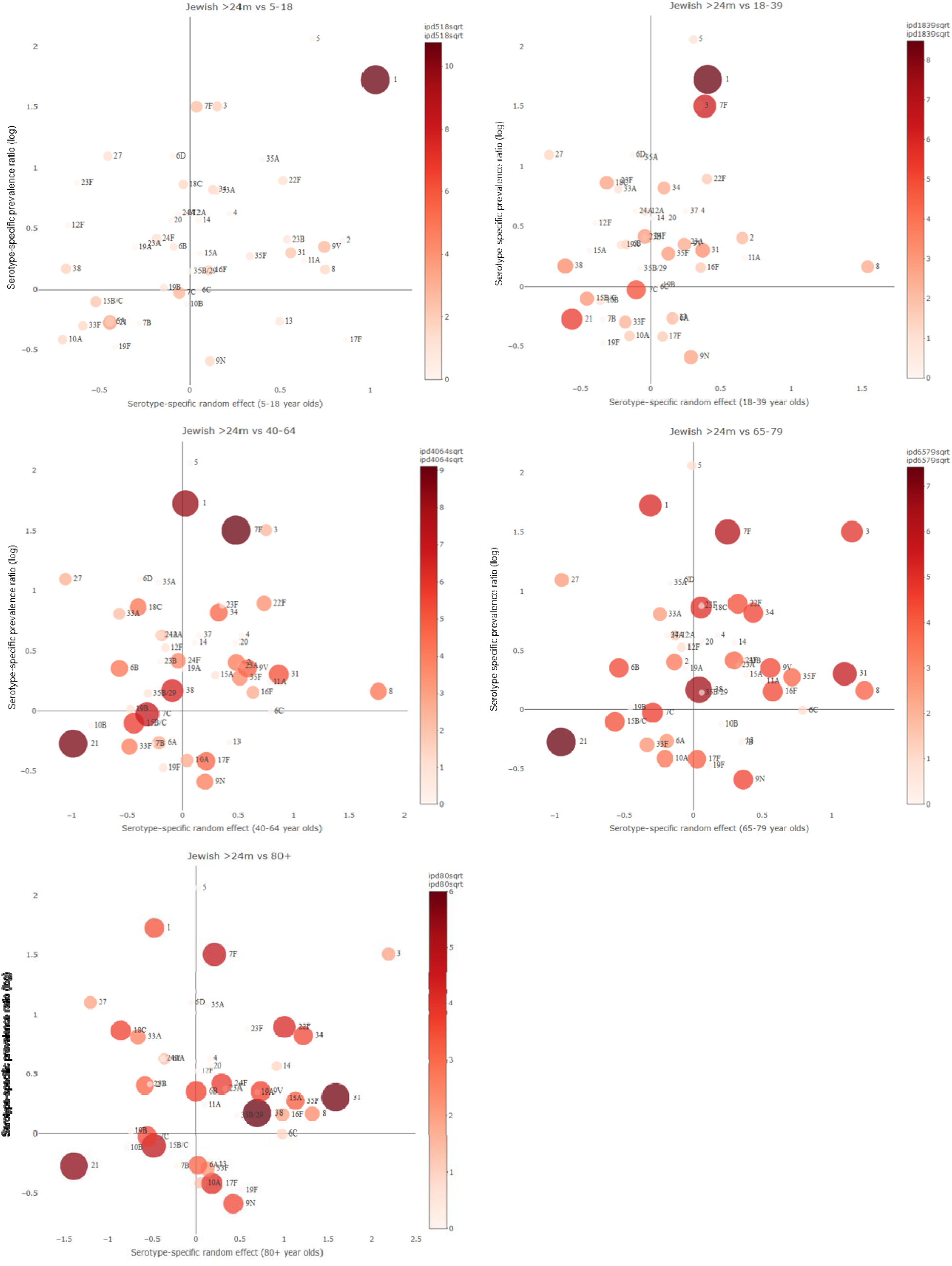
Comparison of serotypes over-represented in IPD in adults with the carriage patterns between older and younger children. The values on the x-axis are the serotype-specific random intercept, which are a measure of how over-represented the serotype is as a cause of IPD in adults. The values on the y-axis denote the serotype-specific log-ratio of prevalence in Jewish children aged >24 months as compared to <24 month olds. Circle size was calculated from the square root of IPD cases, with circle color darkening towards red also denoting higher IPD incidence

While we did not have adult carriage data from Israel, previous studies from the Netherlands have collected carriage data from both children [24] and adults [25]. Notably, there was a correlation between the serotypes that were over-represented as causes of IPD in adults in Israel and the serotypes that were more common among adult carriers compared with pediatric carriers in the Netherlands (**Supplementary Figure S9**). In addition, serotypes that were over-represented as causes of IPD in adults in Israel were also those over-represented as causes of IPD in adults in the Netherlands (as compared to <5 year olds) [10], both prior to and following PCV-implementation (**Supplementary Figure S10**). Interestingly, these correlations were stronger when comparing all adults >65 years of age in the Netherlands to adults aged 65-79 years in Israel than when compared to all adults aged >65 years in Israel, perhaps reflecting some differences between the older age groups in these two countries.

### Serotype-specific patterns of carriage in older children best correlate with IPD in adults

We considered whether carriage prevalence in certain age groups better correlated with IPD in adults, after adjusting for invasiveness. Rather than using arbitrary age groupings, we calculated serotype-specific prevalence for age-bins of different widths. We then evaluated which of these age groupings best correlated with the patterns of serotype-specific IPD in older ages groups. DIC scores obtained from each model demonstrated that serotype patterns in carriage among 36-59 month old children best explained serotype-specific IPD patterns in all of the adult age groups (**Table 2**). Serotype patterns in 24-59 month old children also correlated well with serotype-specific IPD patterns in children and young adults. Patterns were consistent when stratifying by those with or without respiratory complaints.

**Table 2.**
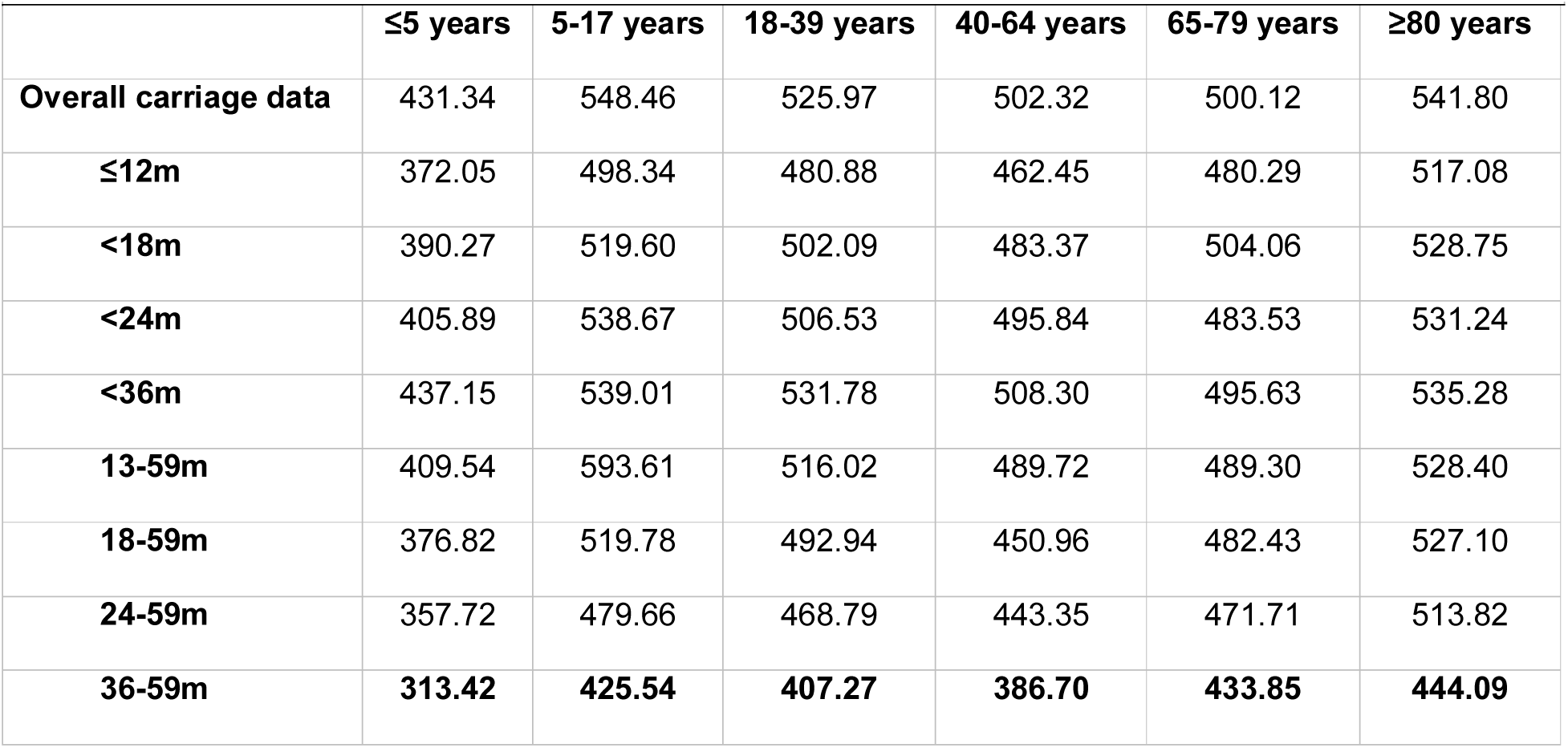
Deviance Information Criteria (DIC) for each model which varied by Jewish child age group

## DISCUSSION

While the serotypes colonizing children are broadly similar around the world, the serotypes causing disease in children and adults vary between age groups and settings [6,26]. Although children <5 years of age are often considered as a single homogenous group when evaluating carriage patterns, our analyses demonstrate that serotype prevalence differs by age. Among the Jewish population in Israel, serotype-specific carriage patterns differ between children <24 months of age and those 24-59 months of age, and the patterns in older children better correlated with patterns of IPD in adults. This finding, along with recent work evaluating the post-vaccine trajectories of carriage and IPD [14], suggests that preschoolers might play a more important role in transmission to adults than infants and toddlers. This has important implications when considering the possible public health impact of different dosing schedules and the timing of booster doses. If older children are important for transmission, then it is critical to ensure that they are protected adequately against colonization.

Importantly, these results highlight that serotype-specific IPD patterns in adults do not always reflect carriage patterns in children (considered the reservoir of pneumococcus in the population). For example, the current high rates of IPD in adults that started in 2015/16 caused by serotypes 8 and 12F were not predicted by carriage surveillance in children in England and Wales [11], nor in the Netherlands [27]. For any strategy aiming to prevent pneumococcal disease, the presence of a large pneumococcal reservoir in older children or adults is an essential factor to consider. Carriage-based surveillance should be designed to address this possibility and to include older children or adults when the goal is to understand or predict indirect effects of PCVs. Further work is needed to understand the implications of these serotype/age patterns for vaccination strategies.

This study focused on the Jewish population in Israel, a population with relatively low transmission of pneumococcus. In this population, the older children attending daycare or preschool have more opportunities for transmission than those at home in isolation (predominantly infants) [13,15]. The situation could differ in populations with higher intensity transmission [28]. Populations with higher intensity transmission tend to have a higher residual burden of vaccine targeted serotypes [28].

The intensity of transmission might also influence the age distribution of serotypes in carriage, and subsequently, in disease. A population with higher intensity transmission would lead to earlier first carriage episodes and more frequent exposure, as has previously been observed in Bedouin versus Jewish children in Israel [29]. An earlier age distribution of carriage in a high transmission setting leads to stronger immunity against the dominant strains (negative frequency dependent selection [30]), potentially allowing the weaker serotypes to colonize older children who are more important for transmission. Conversely, in a low transmission setting, the immunity against the dominant serotypes might be lower, meaning that weaker serotypes are pushed to a later age when general immunity against pneumococcus is high, effectively restraining them within the population. The introduction of vaccines against the dominant serotypes can shift the age distribution of sub-dominant serotypes, influencing disease patterns in adults.

Serotypes over-represented as cause of disease in younger adults were also over-represented in adults considered low risk of pneumococcal disease and *vice versa*, but the patterns differed by serotype. For instance, serotypes 1 and 3 had opposite patterns among adults. Serotype 1 was most over-represented in younger individuals and those without co-morbidities, and serotype 3 was most over-represented in older age groups. Serotype 1 has previously been reported to more commonly affect younger individuals, appearing to have only brief periods of colonization before causing disease, while also primarily infecting previously healthy individuals (in line with younger age) [31]. For this reason, it has been described to act as a “primary pathogen” [31]. When stratifying <18 year olds into smaller age groups, this association with young age was not due to higher rates in 5-10 year olds but rather in the older portion of this group. This points to something specific to the 10-18 year old lifestyle, promoting high circulation, perhaps similar to what is observed for meningococcal disease. With such short periods of colonization, one could expect very few transmission opportunities to parents and even less (and therefore between) elderly individuals (who with younger exposure may also be better protected). In line with this, higher incidence of serotype 1 disease is seen in areas of intense transmission (crowding).

In conclusion, we identified age-related differences in serotype-specific disease, with certain serotypes over- or under-represented in different age groups but also co-morbidity status. These differences are likely explained by differences in susceptibility or exposure to specific serotypes; enhanced carriage data from older children or adults and elderly would help to further understand these patterns. These findings hold importance for those considering new vaccination strategies for infants and for the next generation of adult-specific pneumococcal vaccinations. This issue will gain urgency as current vaccine-serotypes continue to decline and non-vaccine-serotypes increase as cause of disease in adults.

## Data Availability

Due to privacy, raw data analysed in the current study cannot be published. Please contact authors for queries regarding the datasets.

## DECLARATIONS

### Author’s contributions

RD and DMW conceived the study. GRY, NGL, RD and DMW managed the study and collected the data. ALW, JLW, GRY and DMW performed the analyses and interpreted the data. ALW and DMW drafted the manuscript. All authors amended and commented on the final manuscript.

## SUPPLEMENTARY FIGURE LEGENDS

**Supplementary Figure S1**. Pneumococcal carriage prevalence by age for Bedouin (red) and Jewish (blue) children.

**Supplementary Figure S2**. Age distribution of (A) all swabs obtained in the current study for determination of pneumococcal carriage prevalence and (B) for those which tested positive for *Streptococcus pneumoniae*.

**Supplementary Figure S3**. Serotype-specific carriage prevalence in swabs obtained from children with a respiratory-related complaint as compared to those without a respiratory complaint, per study period.

**Supplementary Figure S4**. Serotype-specific carriage prevalence in swabs obtained from Bedouin children as compared to Jewish children, per study period.

**Supplementary Figure S5. Over-representation of serotypes causing IPD in different age groups of adults**. The numbers denote serotype-specific random intercepts from a model fit to IPD data from 80+ year old adults in Israel. Values above zero indicate that the serotype is over-represented as a cause of IPD in this age group based on how frequently they are carried in children <5 years of age and their invasiveness in <5 year olds. Values below zero indicate the serotype is under-represented in IPD. For each serotype the 95% (thinner line) and 68% (thicker line) credible intervals are shown.

**Supplementary Figure S6. Over-representation of serotypes causing IPD for individuals over 18 years of age, stratified according to comorbidity status (no risk, at risk, or high risk for pneumococcal disease) based on recommendations for receipt of PPV23**. The numbers denote serotype-specific random intercepts from a model fit to IPD data from 80+ year old adults in Israel. Values above zero indicate that the serotype is over-represented as a cause of IPD in this age group based on how frequently they are carried in children <5 years of age and their invasiveness in <5 year olds. Values below zero indicate the serotype is under-represented in IPD. For each serotype the 95% (thinner line) and 68% (thicker line) credible intervals are shown.

**Supplementary Figure S7. Heatmap constructed from serotype-specific random intercepts based on carriage and IPD data from Jewish children for individuals aged 18+ years, stratified by comorbidity status (no risk, at risk, or high risk for pneumococcal disease) based on recommendations for receipt of PPV23**. Darker blue represents serotypes in that risk strata under-represented in IPD as expected based on carriage and disease data from children; darker red represents serotypes in that risk strata over-represented in IPD. Serotypes over-represented as cause of disease in younger adults were also over-represented in individuals considered low risk (#). Conversely, serotypes which were over-represented in older adults were over-represented in all risk groups (*). Serotypes under-represented in all adults age groups as compared to based on carriage and disease in children were also under-represented in all risk groups (&).

**Supplementary Figure S8. Heatmap constructed from serotype-specific random intercepts based on carriage and IPD data from Jewish children for all adult age groups over 18 years of age, stratified by both age and co-morbidity status**. Darker blue represents serotypes in that age- and risk-strata under-represented in IPD as expected based on carriage and disease data from children; darker red represents serotypes in that age- and risk-strata over-represented in IPD. Certain serotypes were over-represented as cause of disease in both younger and healthier individuals (#), or just younger individuals regardless of risk status (&). Conversely, other serotypes remained prominent as cause of disease in older age groups, regardless of risk status (*).

**Supplementary Figure S9**. Serotype-specific random effects for adults aged 65 years and older from Israel were compared to the ratio of serotype-specific carriage prevalence in adults over 50 years of age vs that in children under 5 years of age in the Netherlands, both prior to and following the introduction of PCV7 into the Dutch National Immunization Program.

**Supplementary Figure S10**. Serotype-specific random effects for adults over 65 years of age from Israel were compared to the ratio of serotype-specific disease in adults over 50 years of age and children under 5 years of age in the Netherlands, both (A) prior to and (B) following the introduction of PCV7 into the Dutch National Immunization Program. Serotypes over-represented as cause of disease in older adults from Israel were also those over-represented in IPD in older adults in the Netherlands.

## REFERENCES

1. Jansen AGSC, Rodenburg GD, van der Ende A, et al. Invasive pneumococcal disease among adults: associations among serotypes, disease characteristics, and outcome. Clin Infect Dis 2009; 49:e23–9.

2. Wyllie AL, Rümke LW, Arp K, et al. Molecular surveillance on Streptococcus pneumoniae carriage in non-elderly adults; little evidence for pneumococcal circulation independent from the reservoir in children. Sci Rep 2016; 6:34888.

3. Simell B, Auranen K, Käyhty H, Goldblatt D, Dagan R, O’Brien KL. The fundamental link between pneumococcal carriage and disease. Expert Rev Vaccines 2012; 11:841–55.

4. Auranen K, Mehtälä J, Tanskanen A, S Kaltoft M, Kaltoft MS. Between-strain competition in acquisition and clearance of pneumococcal carriage--epidemiologic evidence from a longitudinal study of day-care children. Am J Epidemiol 2010; 171:169–76.

5. Feikin DR, Kagucia EW, Loo JD, et al. Serotype-specific changes in invasive pneumococcal disease after pneumococcal conjugate vaccine introduction: a pooled analysis of multiple surveillance sites. PLoS Med 2013; 10:e1001517.

6. Hausdorff WP, Feikin DR, Klugman KP. Epidemiological differences among pneumococcal serotypes. Lancet Infect Dis 2005; 5:83–93.

7. Weinberger DM, Malley R, Lipsitch M. Serotype replacement in disease after pneumococcal vaccination. Lancet 2011; 378:1962–73.

8. Scott JR, Millar E V, Lipsitch M, et al. Impact of more than a decade of pneumococcal conjugate vaccine use on carriage and invasive potential in Native American communities. J Infect Dis 2012; 205:280–8.

9. Wagenvoort GHJ, Sanders EAM, Vlaminckx BJ, et al. Invasive pneumococcal disease: Clinical outcomes and patient characteristics 2–6 years after introduction of 7-valent pneumococcal conjugate vaccine compared to the pre-vaccine period, the Netherlands. Vaccine 2016; 34:1077–1085.

10. van Deursen AMM, van Mens SP, Sanders EAM, et al. Invasive pneumococcal disease and 7-valent pneumococcal conjugate vaccine, the Netherlands. Emerg Infect Dis 2012; 18:1729–37.

11. Ladhani SN, Collins S, Djennad A, et al. Rapid increase in non-vaccine serotypes causing invasive pneumococcal disease in England and Wales, 2000–17: a prospective national observational cohort study. Lancet Infect Dis 2018; 18:441–451.

12. Flasche S, Van Hoek AJ, Goldblatt D, et al. The Potential for Reducing the Number of Pneumococcal Conjugate Vaccine Doses While Sustaining Herd Immunity in High-Income Countries. PLoS Med 2015; 12:e1001839.

13. Nurhonen M, Cheng AC, Auranen K. Pneumococcal transmission and disease in silico: a microsimulation model of the indirect effects of vaccination. PLoS One 2013; 8:e56079.

14. Weinberger DM, Pitzer VE, Regev-Yochay G, Givon-Lavi N, Dagan R. Association Between the Decline in Pneumococcal Disease in Unimmunized Adults and Vaccine-Derived Protection Against Colonization in Toddlers and Preschool-Aged Children. Am J Epidemiol 2019; 188:160–168.

15. Althouse BM, Hammitt LL, Grant L, et al. Identifying transmission routes of Streptococcus pneumoniae and sources of acquisitions in high transmission communities. Epidemiol Infect 2017; 145:2750–2758.

16. Regev-Yochay G, Paran Y, Bishara J, et al. Early impact of PCV7/PCV13 sequential introduction to the national pediatric immunization plan, on adult invasive pneumococcal disease: A nationwide surveillance study. Vaccine 2015; 33:1135–1142.

17. Ben-Shimol S, Givon-Lavi N, Greenberg D, Dagan R. Pneumococcal nasopharyngeal carriage in children <5 years of age visiting the pediatric emergency room in relation to PCV7 and PCV13 introduction in southern Israel. Hum Vaccines Immunother 2016; 12:268–276.

18. Regev-Yochay G, Katzir M, Strahilevitz J, et al. The herd effects of infant PCV7/PCV13 sequential implementation on adult invasive pneumococcal disease, six years post implementation; a nationwide study in Israel. Vaccine 2017; 35:2449–2456.

19. Weinberger DM, Grant LR, Weatherholtz RC, Warren JL, O’Brien KL, Hammitt LL. Relating Pneumococcal Carriage Among Children to Disease Rates Among Adults Before and After the Introduction of Conjugate Vaccines. Am J Epidemiol 2016; 183:1055–1062.

20. Smith T, Lehmann D, Montgomery J, Gratten M, Riley ID, Alpers MP. Acquisition and invasiveness of different serotypes of Streptococcus pneumoniae in young children. Epidemiol Infect 1993; 111:27–39.

21. Plummer M, Stukalov A, Denwood M. Bayesian Graphical Models using MCMC. 2018.

22. RStudio-Team. RStudio: integrated development for R. RStudio, Inc. 2016.

23. Spiegelhalter DJ, Best NG, Carlin BP, van der Linde A. Bayesian measures of model complexity and fit. J R Stat Soc Ser B (Statistical Methodol 2002; 64:583–639.

24. Wyllie AL, Wijmenga-Monsuur AJ, van Houten MA, et al. Molecular surveillance of nasopharyngeal carriage of Streptococcus pneumoniae in children vaccinated with conjugated polysaccharide pneumococcal vaccines. Sci Rep 2016; 6:23809.

25. Krone CL, Wyllie AL, van Beek J, et al. Carriage of Streptococcus pneumoniae in Aged Adults with Influenza-Like-Illness. PLoS One 2015; 10:e0119875.

26. Hausdorff WP, Bryant J, Paradiso PR, Siber GR. Which pneumococcal serogroups cause the most invasive disease: implications for conjugate vaccine formulation and use, part I. Clin Infect Dis 2000; 30:100–21.

27. Vissers M, Wijmenga-Monsuur AJ, Knol MJ, et al. Increased carriage of non-vaccine serotypes with low invasive disease potential four years after switching to the 10-valent pneumococcal conjugate vaccine in The Netherlands. PLoS One 2018; 13:e0194823.

28. Lourenço J, Obolski U, Swarthout TD, et al. Determinants of high residual post-PCV13 pneumococcal vaccine type carriage in Blantyre, Malawi: a modelling study. bioRxiv 2019; :477695.

29. Lewnard JA, Huppert A, Givon-Lavi N, et al. Density, Serotype Diversity, and Fitness of Streptococcus pneumoniae in Upper Respiratory Tract Cocolonization With Nontypeable Haemophilus influenzae. J Infect Dis 2016; 214:1411–1420.

30. Corander J, Fraser C, Gutmann MU, et al. Frequency-dependent selection in vaccine-associated pneumococcal population dynamics. Nat Ecol Evol 2017; 1:1950–1960.

31. Sjöström K, Spindler C, Ortqvist a, et al. Clonal and capsular types decide whether pneumococci will act as a primary or opportunistic pathogen. Clin Infect Dis 2006; 42:451–9.

